# Combined association of obesity and other cardiometabolic diseases with severe COVID-19 outcomes: a nationwide cross-sectional study of 21,773 Brazilian adult and elderly inpatients

**DOI:** 10.1101/2021.05.14.21257204

**Authors:** Natanael J Silva, Rita C Ribeiro-Silva, Andrêa JF Ferreira, Camila SS Teixeira, Aline S Rocha, Flávia Jôse O Alves, Ila R Falcão, Elizabete J Pinto, Carlos Antônio ST Santos, Rosemeire L Fiaccone, Maria Yury T Ichihara, Enny S Paixão, Mauricio L Barreto

## Abstract

**Objectives:** To investigate the combined association of obesity, diabetes mellitus (DM), and cardiovascular disease (CVD) with severe COVID-19 outcomes in adult and elderly inpatients.

**Design:** Cross-sectional study based on registry data from Brazil’s influenza surveillance system.

**Setting:** Public and private hospitals across Brazil.

**Participants:** Eligible population included 21,942 inpatients aged ≥20 years with positive RT-PCR test for SARS-CoV-2 until Jun 9^th^, 2020.

**Main outcome measures:** Severe COVID-19 outcomes were non-invasive and invasive mechanical ventilation use, ICU admission, and death. Multivariate analyses were conducted separately for adults (20-59 years) and elders (≥60 years) to test the combined association of obesity (without and with DM and/or CVD) and degrees of obesity with each outcome.

**Results:** A sample of 8,848 adults and 12,925 elders were included. Among adults, obesity with DM and/or CVD showed higher prevalence of invasive (PR 3.76, 95%CI 2.82-5.01) and non-invasive mechanical ventilation use (2.06, 1.58-2.69), ICU admission (1.60, 1.40-1.83), and death (1.79, 1.45-2.21) compared with the group without obesity, DM, and CVD. In elders, obesity alone (without DM and CVD) had the highest prevalence of ICU admission (1.40, 1.07-1.82) and death (1.67, 1.00-2.80). In both age groups, obesity alone and combined with DM and/or CVD showed higher prevalence in all outcomes than DM and/or CVD. A dose-response association was observed between obesity and death in adults: class I 1.32 (1.05-1.66), class II 1.41 (1.06-1.87), and class III 1.77 (1.35-2.33).

**Conclusions:** The combined association of obesity, diabetes, and/or CVD with severe COVID-19 outcomes may be stronger in adults than in elders. Obesity alone and combined with DM and/or CVD had more impact on the risk of COVID-19 severity than DM and/or CVD in both age groups. The study also supports an independent relationship of obesity with severe outcomes, including a dose-response association between degrees of obesity and death in adults.

**Article summary:** Strengths and limitations of this study:

- This is the first study that describes the independent and combined relationship of obesity with COVID-19 severity in Brazil, one of the biggest epicenters of the pandemic worldwide.
- The study was based on registry data of a large nationwide sample of patients admitted, due to severe SARS-CoV-2 infection, to public and private hospitals across the country.
- The large sample size and data availability allowed us to analyze the combined association of obesity, diabetes and cardiovascular disease with severe COVID-19 outcomes, separately by age groups and controlled by important confounding variables, e.g. underlying comorbidities.
- The cross-sectional study design does not allow causal inference, and generalization of results must be cautious since only hospitalized cases of severe COVID-19 were included.
- As the study used routinely collected data, which has not been designed primarily for research purposes, it may bring well-known limitations related to missing, underestimation, and potential misclassification.

## Introduction

The coronavirus disease 2019 (COVID-19) pandemic, caused by the severe acute respiratory syndrome coronavirus 2 (SARS-CoV-2), as of 13 May 2021, has already reached more than 160 million infected people and more than 3.3 million deaths in all continents.^1^ Individuals with advanced age and chronic diseases, including cardiometabolic diseases, are considered groups at major risk for complications and severe illness from COVID-19.^2,3^ Obesity has been shown as an independent risk factor for COVID-19 disease.^4–6^ High body mass index (BMI) has been mentioned as a significant risk factor for COVID-19, according to early clinical reports from China,^7^ Italy,^8^ France,^9^ Mexico,^10^ and United States of America.^11^ Several studies have demonstrated that obesity is leading to considerably worse COVID-19 outcomes, especially greater risk of hospital and intensive care unit (ICU) admission, invasive mechanical ventilation, and death.^11–14^

The COVID-19 pandemic is rapidly spreading worldwide, especially in the Americas, where obesity is already a prevalent and important public health problem.^15–16^ Brazil is currently one of the biggest epicenters of the COVID-19 pandemic worldwide, with more than 15.2 million cases and 425 thousand deaths until May 13, 2021.^1^ In 2018, the prevalence of adult overweight and obesity in Brazil was estimated at 55.7% and 19.8%, respectively.^17^ This obesogenic profile of the Brazilian population contributes, among other factors, to the high prevalence of obesity-related diseases such as type 2 diabetes mellitus (DM) and cardiovascular diseases (CVD), in the country.^18^ The fact that individuals with obesity also have more comorbidity diseases, which are either risk factors for COVID-19 severity and death, makes obesity particularly ominous in COVID-19 disease.^10–13^

Several characteristics that can influence the clinical evolution of individuals infected with COVID-19, such as obesity, have been independently documented.^5–6,19^ However, evidence is yet unclear on the combined effect that obesity and obesity-related comorbidities play in COVID-19 severity, especially, in different age groups. We aimed in this study to investigate the combined association of obesity, diabetes, and cardiovascular disease with mechanical ventilation use, ICU admission, and death in a large sample of adult and elderly patients hospitalized with COVID-19 in Brazil. We also explored the independent association between degrees of obesity and the mentioned outcomes.

## Methods

### Study Design and Population

This is a cross-sectional study based on registry data from SIVEP-Gripe (Sistema de Informação de Vigilância Epidemiológica da Gripe), an influenza surveillance system of Brazil’s Ministry of Health. The study used the publicly available dataset of SIVEP-Gripe, which includes de-identified data on cases of severe acute respiratory syndrome across public and private hospitals in Brazil.^20^ These data were obtained through the Rede CoVida’s integrated data platform that has been built with official, open, and authorized data for the production of knowledge about the COVID-19 pandemic. Our study population was composed of patients aged 20 years or older, hospitalized for severe acute respiratory syndrome, with positive RT-PCR test for SARS-CoV-2, and final diagnosis for COVID-19 until Jun 9^th^, 2020. Only cases with complete data on demographic characteristics and comorbidities and plausible BMI values were included in the study.

The study was conducted according to the guidelines laid down in the Declaration of Helsinki. As the study exclusively used publicly available de-identified data, ethics approval by a research ethics committee and informed consent are waived per Resolution n. 466/2012 of the National Health Council of Brazil’s Commission of Ethics in Research.

### Exposure Variable

Obesity was defined as BMI equal to or greater than 30 kg/m^2^, according to the cutoff points proposed by the World Health Organization^21^ and the Pan American Health Organization^22^ for adults and elders, respectively. BMI was calculated by health professionals in the hospital from directly measured or patient self-reported height and weight. Guidelines for the collection and analysis of anthropometric data in health services have been previously standardized by the Ministry of Health.^23^ BMI values <12 or >70 kg/m^2^ were considered implausible and excluded.^24^ Information on the existence of diabetes and any chronic cardiovascular disease was obtained from dichotomous questions (yes/no), which were answered based on patient or family’s report or medical diagnosis.

We created a polytomous four-category variable to evaluate the separate and combined exposure of obesity, diabetes and cardiovascular disease: none/reference (no existence of obesity, diabetes and cardiovascular disease), OB (only existence of obesity), OB + DM and/or CVD (existence of obesity with diabetes and/or cardiovascular disease), and DM and/or CVD (existence of diabetes and/or cardiovascular disease). We also analyzed obesity in adults according to the following degrees of severity based on WHO reference^21^: no obesity (<30 kg/m^2^), obesity class I (≥ 30-34.9 kg/m^2^), obesity class II (≥ 35-39.9 kg/m^2^), and obesity class III (≥ 40 kg/m^2^). Due to the unavailability of BMI cutoff points to classify the degree of obesity in elders, this analysis was only performed for adults.

### Outcome Variables

The severe COVID-19 outcomes were mechanical ventilation use, ICU admission, and death. Information on the use of mechanical ventilation by the patient was obtained and analyzed as a polytomous three-category variable (no use/ use of non-invasive ventilation/ use of invasive ventilation). ICU admission was obtained and analyzed as a dichotomous variable (no/ yes). Death was analyzed as a dichotomous variable based on the patient’s endpoint outcome (cure/ death).

### Covariates

Demographic and comorbidity information were selected as descriptive and confounding variables.^2^ Age in years was calculated from birth and notification dates. Sex was obtained as a dichotomous variable (female/ male). The preexistence of each comorbidity was also obtained as a dichotomous variable (no/ yes): chronic pulmonary disease, asthma, chronic kidney disease, chronic hematologic disease, neurological disease, chronic liver disease, and immunodeficiency/ immunosuppression.

### Statistical Analysis

All analyses were subdivided into adults (≥ 20 and < 60 years) and elders (≥ 60 years). For descriptive analyses, absolute and relative frequencies were calculated for the demographic and comorbidity variables according to the main exposure variable. Multinomial logistic regression models were conducted to test the association of obesity (without and with diabetes and/or CVD) with non-invasive and invasive mechanical ventilation use. To test the association of this exposure variable with ICU admission and death, simple logistic regression models were performed. Same models were analyzed considering the degree of obesity as the main exposure variable for adults. Crude and adjusted estimates were interpreted based on the prevalence ratio (PR) and 95% confidence intervals (95%CI). These estimates were obtained from logistic models using delta method, function ‘prLogisticDelta’, which is implemented in R and available in the package ‘prLogistic’. Adjusted models included the following list of confounding variables: sex, age (years), and the preexistence of chronic pulmonary disease, asthma, kidney disease, hematologic disease, neurological disease, liver disease, and immunodeficiency/ immunosuppression. The models that tested the degrees of obesity were also adjusted for DM and CVD. All analyses were performed using Stata version 15.1 (Stata Corporation, College Station, USA) and R version 3.6.1 (R Foundation for Statistical Computing, Austria).

### Patient and Public Involvement

As the study exclusively used publicly available de-identified data, it was not possible to involve patients or the public in the design, or conduct, or reporting, or dissemination plans of our research.

## Results

During the study period, 21,942 individuals registered in the SIVEP-Gripe were ≥ 20 years old, hospitalized, tested positive for SARS-CoV-2, and had complete demographic and comorbidity information (**Figure 1**). Of these, 169 (0.8%) were excluded due to implausible values of BMI. Of the 21,773 individuals included in the study, 8,848 (40.3%) were adults aged between 20-59 years, and 12,925 (59.6%) were elders aged 60 years or older. Since some patients were still hospitalized on the study endpoint date, information for some outcomes were incomplete. The study samples included in the analysis of each outcome were 8,075 adults and 11,829 elders for mechanical ventilation, 8,414 adults and 12,222 for ICU admission, and 6,565 adults and 9,943 elders for death. Based on demographic and clinical characteristics, the analytical samples in each outcome were very similar to the overall study population and the excluded samples (**Supplementary Table 1**).

**Figure 1.**
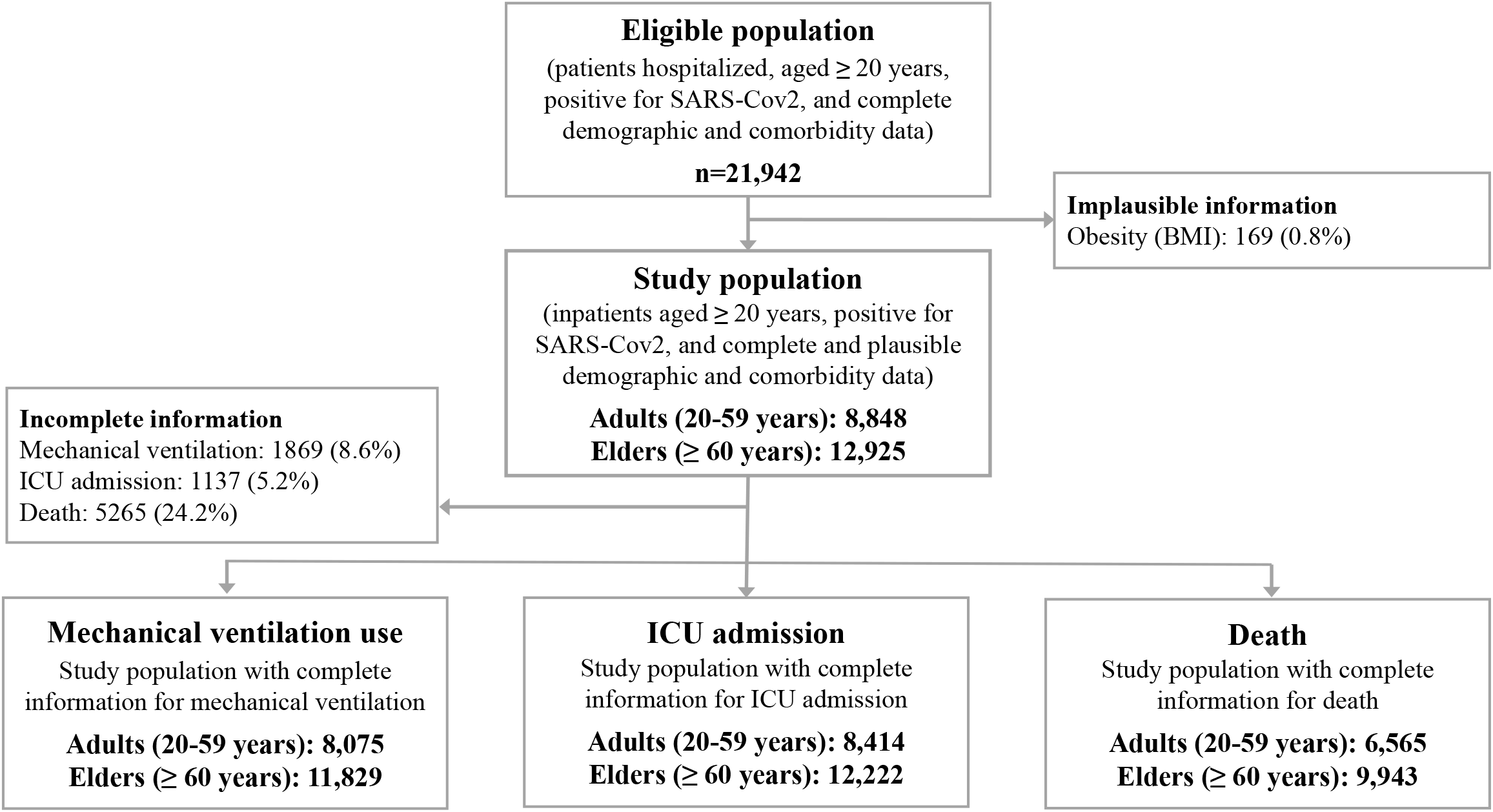
Selection of the study population from SIVEP-Gripe. SIVEP-Gripe: Influenza Epidemiological Surveillance Information System.

The prevalence of obesity was 9.7% in adults and 3.5% in elders. The frequency of obesity without and with DM and/or CVD was respectively 4.6% and 5.1% in adults and 0.7% and 2.8% in elders. Non-invasive and invasive mechanical ventilation were respectively required by 45.0% and 21.2% of adults and 47.0% and 30.0% of elders. ICU admission was needed by 35.4% of adults and 43.6% of elders. Death occurred in 31.1% of adult and 63.0% of elderly patients (**Table 1–2**).

**Table 1.** Demographic characteristics, comorbidities, hospitalization outcomes and death according to the combined exposure of obesity, diabetes, and/or cardiovascular diseases in adults with severe COVID-19.

|  | Total |  | None |  | OB |  | OB + DM<br>and/or CVD |  | DM and/or<br>CVD |  |
| --- | --- | --- | --- | --- | --- | --- | --- | --- | --- | --- |
|  | n | % | n | % | n | % | n | % | n | % |
| Overall | 8848 | 100.0 | 3161 | 35.7 | 409 | 4.6 | 452 | 5.1 | 4826 | 54.6 |
| **Sex** |  |  |  |  |  |  |  |  |  |  |
| Female | 3774 | 42.7 | 1511 | 40.0 | 165 | 4.4 | 199 | 5.3 | 1899 | 50.3 |
| Male | 5074 | 57.4 | 1650 | 32.5 | 244 | 4.8 | 253 | 5.0 | 2927 | 57.7 |
| **Age** |  |  |  |  |  |  |  |  |  |  |
| < 40 years | 1976 | 22.3 | 1064 | 53.9 | 188 | 9.5 | 102 | 5.2 | 622 | 31.5 |
| ≥ 40 years | 6872 | 77.7 | 2097 | 30.5 | 221 | 3.2 | 350 | 5.1 | 4204 | 61.2 |
| **Chronic pulmonary disease** |  |  |  |  |  |  |  |  |  |  |
| No | 8502 | 96.1 | 2969 | 34.9 | 388 | 4.6 | 435 | 5.1 | 4710 | 55.4 |
| Yes | 346 | 3.9 | 192 | 55.5 | 21 | 6.1 | 17 | 4.9 | 116 | 33.5 |
| **Asthma** |  |  |  |  |  |  |  |  |  |  |
| No | 8184 | 92.5 | 2728 | 33.3 | 383 | 4.7 | 414 | 5.1 | 4659 | 56.9 |
| Yes | 664 | 7.5 | 433 | 65.2 | 26 | 3.9 | 38 | 5.7 | 167 | 25.2 |
| **Chronic kidney disease** |  |  |  |  |  |  |  |  |  |  |
| No | 8297 | 93.8 | 2958 | 35.7 | 399 | 4.8 | 434 | 5.2 | 4506 | 54.3 |
| Yes | 551 | 6.2 | 203 | 36.8 | 10 | 1.8 | 18 | 3.3 | 320 | 58.1 |
| **Chronic hematologic disease** |  |  |  |  |  |  |  |  |  |  |
| No | 8710 | 98.4 | 3081 | 35.4 | 406 | 4.7 | 445 | 5.1 | 4778 | 54.9 |
| Yes | 138 | 1.6 | 80 | 58.0 | 3 | 2.2 | 7 | 5.1 | 48 | 34.8 |
| **Chronic neurological disease** |  |  |  |  |  |  |  |  |  |  |
| No | 8588 | 97.1 | 3014 | 35.1 | 406 | 4.7 | 442 | 5.2 | 4726 | 55.0 |
| Yes | 260 | 2.9 | 147 | 56.5 | 3 | 1.2 | 10 | 3.9 | 100 | 38.5 |
| **Chronic liver disease** |  |  |  |  |  |  |  |  |  |  |
| No | 8684 | 98.2 | 3083 | 35.5 | 406 | 4.7 | 443 | 5.1 | 4752 | 54.7 |
| Yes | 164 | 1.9 | 78 | 47.6 | 3 | 1.8 | 9 | 5.5 | 74 | 45.1 |
| **Immunosuppression** |  |  |  |  |  |  |  |  |  |  |
| No | 8276 | 93.5 | 2777 | 33.6 | 393 | 4.8 | 440 | 5.3 | 4666 | 56.4 |
| Yes | 572 | 6.5 | 384 | 67.1 | 16 | 2.8 | 12 | 2.1 | 160 | 28.0 |
| **Mechanical ventilation\*** |  |  |  |  |  |  |  |  |  |  |
| No | 2727 | 33.8 | 1144 | 42.0 | 93 | 3.4 | 88 | 3.2 | 1402 | 51.4 |
| Non-invasive | 3634 | 45.0 | 1178 | 32.4 | 192 | 5.3 | 190 | 5.2 | 2074 | 57.1 |
| Invasive | 1714 | 21.2 | 529 | 30.9 | 101 | 5.9 | 150 | 8.8 | 934 | 54.5 |
| **ICU admission\*** |  |  |  |  |  |  |  |  |  |  |
| No | 5438 | 64.6 | 2025 | 37.2 | 235 | 4.3 | 222 | 4.1 | 2956 | 54.4 |
| Yes | 2976 | 35.4 | 1007 | 33.8 | 163 | 5.5 | 216 | 7.3 | 1590 | 53.4 |
| **Death\*** |  |  |  |  |  |  |  |  |  |  |
| No | 4525 | 68.9 | 1699 | 37.6 | 211 | 4.7 | 200 | 4.4 | 2415 | 53.4 |
| Yes | 2040 | 31.1 | 640 | 31.4 | 92 | 4.5 | 140 | 6.9 | 1168 | 57.3 |

OB: obesity (BMI≥30 kg/m<sup>2</sup>), DM: diabetes mellitus, CVD: cardiovascular disease, ICU: intensive care unit.
\* Mechanical ventilation (n=8075), ICU admission (n=8414), and death (n=6565).

**Table 2.**
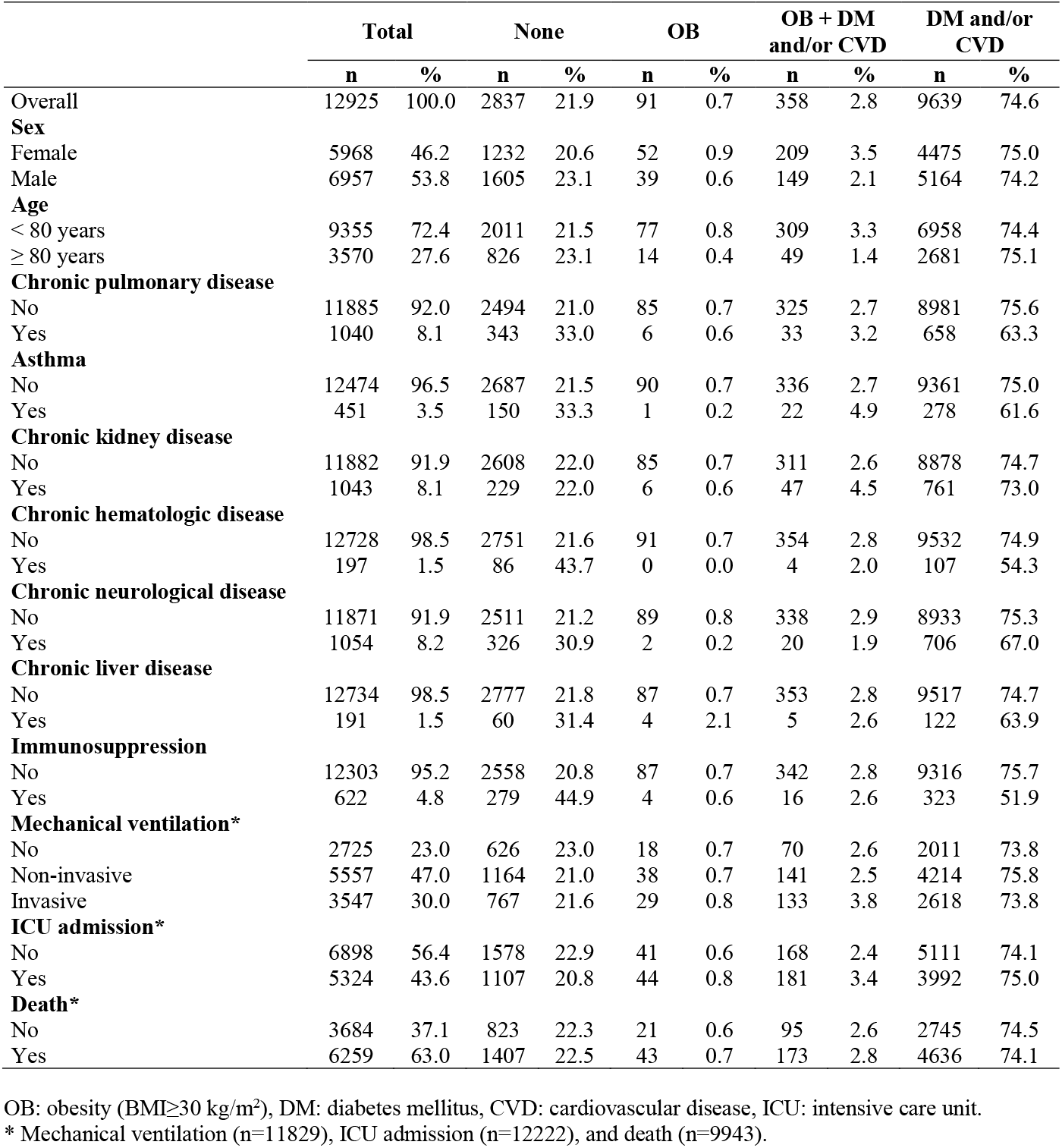
Demographic characteristics, comorbidities, hospitalization outcomes and death according to the combined exposure of obesity, diabetes, and/or cardiovascular diseases in elders with severe COVID-19.

|  | Total |  | None |  | OB |  | OB + DM<br>and/or CVD |  | DM and/or<br>CVD |  |
| --- | --- | --- | --- | --- | --- | --- | --- | --- | --- | --- |
|  | n | % | n | % | n | % | n | % | n | % |
| Overall | 12925 | 100.0 | 2837 | 21.9 | 91 | 0.7 | 358 | 2.8 | 9639 | 74.6 |
| **Sex** |  |  |  |  |  |  |  |  |  |  |
| Female | 5968 | 46.2 | 1232 | 20.6 | 52 | 0.9 | 209 | 3.5 | 4475 | 75.0 |
| Male | 6957 | 53.8 | 1605 | 23.1 | 39 | 0.6 | 149 | 2.1 | 5164 | 74.2 |
| **Age** |  |  |  |  |  |  |  |  |  |  |
| < 80 years | 9355 | 72.4 | 2011 | 21.5 | 77 | 0.8 | 309 | 3.3 | 6958 | 74.4 |
| ≥ 80 years | 3570 | 27.6 | 826 | 23.1 | 14 | 0.4 | 49 | 1.4 | 2681 | 75.1 |
| **Chronic pulmonary disease** |  |  |  |  |  |  |  |  |  |  |
| No | 11885 | 92.0 | 2494 | 21.0 | 85 | 0.7 | 325 | 2.7 | 8981 | 75.6 |
| Yes | 1040 | 8.1 | 343 | 33.0 | 6 | 0.6 | 33 | 3.2 | 658 | 63.3 |
| **Asthma** |  |  |  |  |  |  |  |  |  |  |
| No | 12474 | 96.5 | 2687 | 21.5 | 90 | 0.7 | 336 | 2.7 | 9361 | 75.0 |
| Yes | 451 | 3.5 | 150 | 33.3 | 1 | 0.2 | 22 | 4.9 | 278 | 61.6 |
| **Chronic kidney disease** |  |  |  |  |  |  |  |  |  |  |
| No | 11882 | 91.9 | 2608 | 22.0 | 85 | 0.7 | 311 | 2.6 | 8878 | 74.7 |
| Yes | 1043 | 8.1 | 229 | 22.0 | 6 | 0.6 | 47 | 4.5 | 761 | 73.0 |
| **Chronic hematologic disease** |  |  |  |  |  |  |  |  |  |  |
| No | 12728 | 98.5 | 2751 | 21.6 | 91 | 0.7 | 354 | 2.8 | 9532 | 74.9 |
| Yes | 197 | 1.5 | 86 | 43.7 | 0 | 0.0 | 4 | 2.0 | 107 | 54.3 |
| **Chronic neurological disease** |  |  |  |  |  |  |  |  |  |  |
| No | 11871 | 91.9 | 2511 | 21.2 | 89 | 0.8 | 338 | 2.9 | 8933 | 75.3 |
| Yes | 1054 | 8.2 | 326 | 30.9 | 2 | 0.2 | 20 | 1.9 | 706 | 67.0 |
| **Chronic liver disease** |  |  |  |  |  |  |  |  |  |  |
| No | 12734 | 98.5 | 2777 | 21.8 | 87 | 0.7 | 353 | 2.8 | 9517 | 74.7 |
| Yes | 191 | 1.5 | 60 | 31.4 | 4 | 2.1 | 5 | 2.6 | 122 | 63.9 |
| **Immunosuppression** |  |  |  |  |  |  |  |  |  |  |
| No | 12303 | 95.2 | 2558 | 20.8 | 87 | 0.7 | 342 | 2.8 | 9316 | 75.7 |
| Yes | 622 | 4.8 | 279 | 44.9 | 4 | 0.6 | 16 | 2.6 | 323 | 51.9 |
| **Mechanical ventilation\*** |  |  |  |  |  |  |  |  |  |  |
| No | 2725 | 23.0 | 626 | 23.0 | 18 | 0.7 | 70 | 2.6 | 2011 | 73.8 |
| Non-invasive | 5557 | 47.0 | 1164 | 21.0 | 38 | 0.7 | 141 | 2.5 | 4214 | 75.8 |
| Invasive | 3547 | 30.0 | 767 | 21.6 | 29 | 0.8 | 133 | 3.8 | 2618 | 73.8 |
| **ICU admission\*** |  |  |  |  |  |  |  |  |  |  |
| No | 6898 | 56.4 | 1578 | 22.9 | 41 | 0.6 | 168 | 2.4 | 5111 | 74.1 |
| Yes | 5324 | 43.6 | 1107 | 20.8 | 44 | 0.8 | 181 | 3.4 | 3992 | 75.0 |
| **Death\*** |  |  |  |  |  |  |  |  |  |  |
| No | 3684 | 37.1 | 823 | 22.3 | 21 | 0.6 | 95 | 2.6 | 2745 | 74.5 |
| Yes | 6259 | 63.0 | 1407 | 22.5 | 43 | 0.7 | 173 | 2.8 | 4636 | 74.1 |

OB: obesity (BMI≥30 kg/m<sup>2</sup>), DM: diabetes mellitus, CVD: cardiovascular disease, ICU: intensive care unit.
\* Mechanical ventilation (n=11829), ICU admission (n=12222), and death (n=9943).

In the adjusted analyses for adults, obesity alone (without DM and CVD) was associated with an increased prevalence of invasive (PR 2.69, 95%CI 1.98-3.65) and non-invasive mechanical ventilation need (PR 2.13, 95%CI 1.64-2.78), ICU admission (PR 1.31, 95%CI 1.13-1.53), and death (PR 1.33, 95%CI 1.05-1.69) when compared with the group without obesity, DM, and CVD. Obesity with DM and/or CVD was associated with an even higher prevalence of invasive mechanical ventilation (PR 3.76, 95%CI 2.82-5.01) and non-invasive ventilation use (PR 2.06, 95%CI 1.58-2.69), ICU admission (PR 1.60, 95%CI 1.40-1.83), and death in adults (PR 1.79, 95%CI 1.45-2.21). The subgroup of adults with DM and/or CVD showed in general the lowest prevalence ratios for all analyzed outcomes than the subgroups with the presence of obesity alone or combined (**Table 3**).

**Table 3.** Combined association of obesity, diabetes, and/or cardiovascular disease with non-invasive and invasive mechanical ventilation use, intensive care unit admission, and death in adult and elderly patients hospitalized with severe COVID-19.

| Main exposure variable |  | Non-invasive mechanical ventilation* |  |  |  | Invasive mechanical ventilation* |  |  |  |
| --- | --- | --- | --- | --- | --- | --- | --- | --- | --- |
|  |  | Crude model |  | Adjusted model # |  | Crude model |  | Adjusted model # |  |
|  |  | PR | 95% CI | PR | 95% CI | PR | 95% CI | PR | 95% CI |
| **Adults**<br>20-59 years | None | 1.00 |  | 1.00 |  | 1.00 |  | 1.00 |  |
|  | OB | 2.00 | 1.54-2.60 | 2.13 | 1.64-2.78 | 2.35 | 1.74-3.17 | 2.69 | 1.98-3.65 |
|  | OB + DM and/or CVD | 2.10 | 1.61-2.73 | 2.06 | 1.58-2.69 | 3.69 | 2.78-4.89 | 3.76 | 2.82-5.01 |
|  | DM and/or CVD | 1.44 | 1.29-1.60 | 1.35 | 1.20-1.51 | 1.44 | 1.26-1.64 | 1.32 | 1.14-1.52 |
| **Elders**<br>≥ 60 years | None | 1.00 |  | 1.00 |  | 1.00 |  | 1.00 |  |
|  | OB | 1.14 | 0.64-2.01 | 1.22 | 0.69-2.16 | 1.31 | 0.72-2.39 | 1.43 | 0.78-2.61 |
|  | OB + DM and/or CVD | 1.08 | 0.80-1.47 | 1.15 | 0.84-1.55 | 1.55 | 1.14-2.11 | 1.66 | 1.22-2.27 |
|  | DM and/or CVD | 1.13 | 1.01-1.26 | 1.14 | 1.01-1.27 | 1.06 | 0.94-1.20 | 1.10 | 0.97-1.24 |
|  |  | ICU admission** |  |  |  | Death*** |  |  |  |
|  |  | Crude model |  | Adjusted model # |  | Crude model |  | Adjusted model # |  |
|  |  | PR | 95% CI | PR | 95% CI | PR | 95% CI | PR | 95% CI |
| **Adults**<br>20-59 years | None | 1.00 |  | 1.00 |  | 1.00 |  | 1.00 |  |
|  | OB | 1.23 | 1.08-1.40 | 1.31 | 1.13-1.53 | 1.11 | 0.92-1.33 | 1.33 | 1.05-1.69 |
|  | OB + DM and/or CVD | 1.48 | 1.33-1.65 | 1.60 | 1.40-1.83 | 1.50 | 1.30-1.74 | 1.79 | 1.45-2.21 |
|  | DM and/or CVD | 1.05 | 0.99-1.12 | 1.03 | 0.95-1.12 | 1.19 | 1.10-1.29 | 1.16 | 1.03-1.30 |
| **Elders**<br>≥ 60 years | None | 1.00 |  | 1.00 |  | 1.00 |  | 1.00 |  |
|  | OB | 1.26 | 1.02-1.55 | 1.40 | 1.07-1.82 | 1.06 | 0.89-1.27 | 1.67 | 1.00-2.80 |
|  | OB + DM and/or CVD | 1.26 | 1.13-1.41 | 1.37 | 1.19-1.59 | 1.02 | 0.93-1.12 | 1.39 | 1.07-1.80 |
|  | DM and/or CVD | 1.06 | 1.01-1.12 | 1.11 | 1.04-1.18 | 1.00 | 0.96-1.03 | 1.05 | 0.95-1.16 |

OB: obesity (BMI ≥ 30 kg/m<sup>2</sup>), DM: diabetes mellitus, CVD: cardiovascular disease, ICU: intensive care unit, PR: prevalence ratio, 95% CI: 95% confidence interval.
\* Crude and adjusted multinomial logistic regression models for mechanical ventilation use in adults (n=8075) and elders (n=11829).
\*\* Crude and adjusted logistic regression models for ICU admission in adults (n= 8414) and elders (n=12222).
\*\*\* Crude and adjusted logistic regression models for death in adults (n=6565) and elders (n=9943).

Among elders, obesity without DM and CVD increased independently the prevalence of ICU admission by 40% (95%CI 1.07-1.82) and death by 67% (1.00-2.80). To a lesser extent, obesity with DM and/or CVD was also associated with an increased prevalence of invasive mechanical ventilation need (PR 1.66, 95%CI 1.22-2.27), ICU admission (PR 1.37, 95%CI 1.19-1.59), and death (PR 1.39, 95%CI 1.07-1.80). Elders with DM and/or CVD had the lowest prevalence ratios for the analyzed outcomes than the subgroups of elders with obesity alone or combined (**Table 3**). In the analyses by the degree of obesity, we did not observe much difference in the prevalence of adverse outcomes, except for the prevalence of death that increased with the severity of obesity: Class I 1.32 (95%CI 1.05-1.66), Class II 1.41 (1.06-1.87), and Class III 1.77 (1.35-2.33) (**Table 4**).

**Table 4.** Independent association of degrees of obesity with non-invasive and invasive mechanical ventilation, intensive care unit admission, and death in hospitalized adults with severe COVID-19.

| Main exposure variable | Non-invasive mechanical ventilation* |  |  |  | Invasive mechanical ventilation* |  |  |  |
| --- | --- | --- | --- | --- | --- | --- | --- | --- |
|  | Crude model |  | Adjusted model # |  | Crude model |  | Adjusted model # |  |
|  | PR | 95% CI | PR | 95% CI | PR | 95% CI | PR | 95% CI |
| No obesity (< 30 kg/m<sup>2</sup>) | 1.00 |  | 1.00 |  | 1.00 |  | 1.00 |  |
| Obesity class I (≥ 30-34.9 kg/m<sup>2</sup>) | 1.78 | 1.35-2.33 | 1.91 | 1.45-2.51 | 2.59 | 1.93-3.47 | 3.00 | 2.22-4.05 |
| Obesity class II (≥ 35-39.9 kg/m<sup>2</sup>) | 1.44 | 1.04-2.00 | 1.58 | 1.14-2.19 | 2.10 | 1.47-2.99 | 2.47 | 1.72-3.54 |
| Obesity class III (≥ 40 kg/m<sup>2</sup>) | 1.70 | 1.19-2.44 | 1.88 | 1.31-2.69 | 2.51 | 1.71-3.70 | 3.00 | 2.03-4.45 |
|  | ICU admission** |  |  |  | Death** |  |  |  |
|  | Crude model |  | Adjusted model # |  | Crude model |  | Adjusted model # |  |
|  | PR | 95% CI | PR | 95% CI | PR | 95% CI | PR | 95% CI |
| No obesity (< 30 kg/m<sup>2</sup>) | 1.00 |  | 1.00 |  | 1.00 |  | 1.00 |  |
| Obesity class I (≥ 30-34.9 kg/m<sup>2</sup>) | 1.31 | 1.17-1.47 | 1.42 | 1.23-1.64 | 1.11 | 0.94-1.31 | 1.32 | 1.05-1.66 |
| Obesity class II (≥ 35-39.9 kg/m<sup>2</sup>) | 1.34 | 1.16-1.54 | 1.46 | 1.23-1.74 | 1.16 | 0.95-1.42 | 1.41 | 1.06-1.87 |
| Obesity class III (≥ 40 kg/m<sup>2</sup>) | 1.32 | 1.14-1.54 | 1.45 | 1.20-1.74 | 1.33 | 1.10-1.59 | 1.77 | 1.35-2.33 |

Degrees of obesity defined by the WHO cutoff points.
PR: prevalence ratio, 95%CI: 95% confidence interval.
\* Crude and adjusted multinomial logistic regression models for mechanical ventilation use (n=8075).
\*\* Crude and adjusted logistic regression models for ICU admission (n=8414) and mortality (n=6565).

## Discussion

This is the first study that describe the relationship of obesity and COVID-19 in Brazil, based on a large nationwide sample of adults and elders tested positive for SARS-CoV-2 and admitted to public and private hospitals. Our results highlights that obesity with DM and/or CVD was associated with higher rates of invasive mechanical ventilation use, ICU admission, and death in adults, while obesity alone (without DM and CVD) was associated with higher rates of ICU admission and death among elders. In both age groups, obesity alone and obesity combined with DM and/or CVD had more impact on the risk of all severe COVID-19 outcomes than the subgroup with DM and/or CVD. The study also supports the independent association of obesity with the analyzed outcomes and a dose-response association between degrees of obesity and death in adults.

Some mechanisms related to the role of obesity and related diseases in worsening the clinical condition of patients affected by SARS-CoV-2 have been pointed out: i) greater body weight causes less elasticity of the chest wall and less total compliance of the respiratory system, leading to a restriction of the ventilation and the excursion of the diaphragm, making difficult the airway management in patients with obesity;^25^ ii) obesity is associated with sleep apnea syndrome and chronic obstructive pulmonary disease, which lead to surfactant dysfunction and impede the proper functioning of the airways;^26^ iii) obesity is a metabolic and inflammatory disease, which is associated with the development or worsening of other chronic and endocrine comorbidities (e.g. type 2 diabetes, hypertension, dyslipidemia and CVD) that can modify innate and adaptive immune responses, making the immune system more vulnerable to infections and less responsive to antivirals and antimicrobial drugs;^16^ iv) glycemic decompensation, common in patients with obesity, is associated with impaired ventilation function.^26^

It is important to note that the COVID-19 pandemic imposes a double burden of disease, especially among the elderly individuals, since the prevalence of diabetes, hypertension, cardiovascular diseases, and other comorbidities associated with COVID-19 severity increases with age.^3,27^ However, our study suggests that obesity combined with diabetes and/or cardiovascular disease may offer higher risk of COVID-19 severity for adults although the overall prevalence of diseases and rates of ICU admission and mortality were higher in elders. Obesity alone seemed to provide higher risk of severe outcomes, especially death, in elders.

Few studies to-date have explored the combined and additional effect of obesity on COVID-19 severity.^13,28^ A study investigated the patterns of multimorbidity among fatal cases of COVID-19 in Colombia.^28^ Similar to our study, the authors found that obesity alone or with other diseases was associated with a higher risk of COVID-19 fatality among young people. Furthermore, a population-based study in Mexico observed that the addition of obesity to any number of comorbidities significantly increased the risk of COVID-19 lethality.^13^ Using a causally ordered mediation analysis, this study also found that 49.5% of the effect of diabetes on COVID-19 lethality was mediated by obesity, particularly in early-onset cases < 40 years of age.

Other studies also suggest that obesity is independently associated with severe outcomes of COVID-19, regardless of age and other associated comorbidities.^11–14^ A large study in Mexico^13^ showed that patients with obesity had higher rates of ICU admission and were more likely to be intubated in relation to patients without obesity. This study also found a five-fold increased risk of mortality due to COVID-19 in patients with obesity.^13^ In a hospital-based study in France, it was observed that BMI > 35 kg/m^2^ was associated with the need for invasive mechanical ventilation.^14^ Few studies to-date have similarly found a dose-response association between degrees of obesity and COVID-19 death.^29^ Based on care records of 17,278,392 UK adults, the study showed that the risk of COVID-19 death increases independently with the degree of obesity: 30-34.9 kg/m^2^ (HR 1.05), 35-39.9 kg/m^2^ (1.40), and ≥40 kg/m^2^ (2.66).^29^ Other studies have evidenced the association of obesity with COVID-19 complications and death among adults.^12,30^ A hospital-based study in New York City showed that morbid obesity (BMI ≥ 40 kg/m^2^) is strongly and independently associated with death in hospitalized patients younger than 50 years.^30^ Another study in New York City found a similar dose-response relationship between degrees of obesity and acute and critical care.^12^ Patients less than 60 years old with BMI between 30 and 34.9 kg/m^2^ (obesity class I) were 2.0 and 1.8 times more likely to be respectively admitted for acute care (general hospital admission) and critical care (ICU admission or invasive ventilator) compared to individuals with BMI < 30 kg/m^2^. Patients of the same age group with BMI ≥ 35 kg/m^2^ (obesity class II and III) showed 2.2 and 3.6 more chances of being hospitalized for acute and critical care, respectively.^12^

### Strengths and limitations

One of the greatest strengths of the study was the use of SIVEP-Gripe dataset. Because severe acute respiratory syndrome is a condition of compulsory notification in both public and private hospitals,^31^ we have a nationwide representative sample of patients hospitalized for severe COVID-19 in Brazil. In addition, the large sample sizes allowed us to analyze adults and elders separately, as well as the degrees of obesity which dose-response association with death was evidenced. The availability of important confounding variables (sex, age, and preexisting comorbidities) to control the estimated associations, as well as hospital outcomes and mortality of COVID-19, was another differential of the study. Only patients with positive RT-PCR test for SARS-CoV-2 and final diagnosis for COVID-19 were included which gives greater precision on the studied population. The availability and use of data from health surveillance systems may be a lesson from Brazil that other countries can learn for obtaining routine and timely data to guide health systems and research in preparing and responding to pandemics before and during their course.

The study also has some limitations that must be considered. Because this is a cross-sectional study, a causal association cannot be inferred. As we used routinely collected data, which has not been designed primarily for research purposes, it may bring well-known limitations related to missing, underestimation, and potential misclassification. Obesity prevalence may have been underestimated due to the completeness of obesity and BMI data. Previous studies using SIVEP-Gripe data have also found a low prevalence of obesity in this population.^32,33^ Better routine collection of height and weight data is still needed in clinical practice. Also, we believe that health professionals have adopted more the one method to collect weight and height information for BMI calculation, such as the patient’s self-report and direct measure. Therefore, in addition to BMI which implausible values were checked and excluded, the classification of obesity was also confirmed from a dichotomous variable on the presence of obesity (no/yes). Although it is known that BMI does not distinguish between fat and lean body mass, and thus may lead to misclassification bias, BMI has been shown as a strong predictor of excess body fat and has been widely used in epidemiological studies.^15^ Information for some outcomes were incomplete because some patients were still hospitalized on the study endpoint date. However, that did not represent a potential selection bias to our study. The analytical samples in each outcome had similar demographic and clinical characteristics than the overall study population and the excluded samples (Supplementary Table 1). Data on ethnicity/race was very incomplete, and thus was not included in the analysis. Additional studies are needed to further explore the relationship between socioeconomic characteristics and obesity in severe disease. Finally, the generalization of results must be cautious since the study included only hospitalized cases of COVID-19.

## Conclusions

The combined association of obesity, diabetes, and/or cardiovascular disease with severe COVID-19 outcomes, especially ICU admission and death, may be stronger in adult than in elderly inpatients. In both age groups, obesity alone and obesity combined with DM and/or CVD had more impact on the risk of all severe COVID-19 outcomes than the subgroup with DM and/or CVD. The study also supports an independent relationship of obesity with the severe outcomes, including a dose-response association between degrees of obesity and death in adults. These findings suggest important implications for the clinical care of patients with obesity and severe COVID-19, such as the increased need of critical care and higher risk of death among these patients. Our study also supports the inclusion of people with obesity, independently of other preexisting comorbidities and age, in the high-risk and vaccine priority groups for protection from SARS-CoV-2 infection.

## Data Availability

Data is freely available without restriction at https://opendatasus.saude.gov.br/dataset/bd-srag-2020. Code book and analytic code will be made available upon request from the corresponding author.

## Acknowledgments

The authors thank the members of Rede CoVida’s Epidemiology & Information Group for the work of identifying and collecting data related to COVID-19.

## Author Contributions

NJS, RCRS, and RLF designed the study and analysis strategy. NJS, CASTS, and MYTI obtained, documented, and described the data. AJFF, CSST, ASR, FJOA, and IRF carried out the literature search. NJS and EJP performed the data analysis. NJS, RCRS, AJFF, CSST, ASR, FJOA, IRF, ESP and MLB contributed to data interpretation. NJS, AJFF, CSST, ASR, FJOA, and IRF drafted the manuscript. RCRS, ESP, MYTI, and MLB critically revised the manuscript. All authors read and approved the final manuscript.

## Patient Consent for Publication

Not required.

## References

1. World Health Organization. WHO Coronavirus Disease (COVID-19) Dashboard. https://covid19.who.int/. Accessed May 13, 2020.

2. Centers for Disease Control and Prevention. Coronavirus Disease 2019 (COVID-19). People Who Are at Increased Risk for Severe Illness. https://www.cdc.gov/coronavirus/2019-ncov/need-extra-precautions/people-at-increased-risk.html. Accessed July 21, 2020.

3. Liu H, Chen S, Liu M, Nie H, Lu H. Comorbid Chronic Diseases are Strongly Correlated with Disease Severity among COVID-19 Patients: A Systematic Review and Meta-Analysis. Aging Dis. 2020;11(3):668–678. doi:10.14336/ad.2020.0502.

4. Alberca RW, Oliveira LM, Branco ACCC, Pereira NZ, Sato MN. Obesity as a risk factor for COVID-19: an overview [published online June 15, 2020]. Crit Rev Food Sci Nutr. doi:10.1080/10408398.2020.1775546.

5. Sattar N, McInnes IB, McMurray JJV. Obesity Is a Risk Factor for Severe COVID-19 Infection: Multiple Potential Mechanisms. Circulation. 2020;142(1):4–6. doi:10.1161/circulationaha.120.047659.

6. Caussy C, Wallet F, Laville M, Disse E. Obesity is Associated with Severe Forms of COVID-19. Obesity (Silver Spring). 2020;28(7):1175. doi:10.1002/oby.22842.

7. Li Q, Guan X, Wu P, et al. Early Transmission Dynamics in Wuhan, China, of Novel Coronavirus-Infected Pneumonia. N Engl J Med. 2020;382(13):1199–1207. doi:10.1056/nejmoa2001316.

8. Grasselli G, Zangrillo A, Zanella A, et al. Baseline Characteristics and Outcomes of 1591 Patients Infected with SARS-CoV-2 Admitted to ICUs of the Lombardy Region, Italy. JAMA. 2020;323(16):1574–1581. doi:10.1001/jama.2020.5394.

9. Caussy C, Pattou F, Wallet F, et al. Prevalence of obesity among adult inpatients with COVID-19 in France. Lancet Diabetes Endocrinol. 2020;8(7):562–564. doi:10.1016/s2213-8587(20)30160-1.

10. Hernández-Garduño E. Obesity is the comorbidity more strongly associated for Covid-19 in Mexico. A case-control study. Obes Res Clin Pract. 2020;14(4):375–379. doi:10.1016/j.orcp.2020.06.001.

11. Hajifathalian K, Kumar S, Newberry C, et al. Obesity is associated with worse outcomes in COVID-19: Analysis of Early Data From New York City. Obesity (Silver Spring). 2020;28(9):1606–1612. doi:10.1002/oby.22923.

12. Lighter J, Phillips M, Hochman S, et al. Obesity in patients younger than 60 years is a risk factor for Covid-19 hospital admission. Clin Infect Dis. 2020;71(15):896–897. doi:10.1093/cid/ciaa415.

13. Bello-Chavolla OY, Bahena-López JP, Antonio-Villa NE, et al. Predicting Mortality Due to SARS-CoV-2: A Mechanistic Score Relating Obesity and Diabetes to COVID-19 Outcomes in Mexico. J Clin Endocrinol Metab. 2020;105(8):dgaa346. doi:10.1210/clinem/dgaa346.

14. Simonnet A, Chetboun M, Poissy J, et al. High Prevalence of Obesity in Severe Acute Respiratory Syndrome Coronavirus-2 (SARS-CoV-2) Requiring Invasive Mechanical Ventilation. Obesity (Silver Spring). 2020;28(7):1195–1199. doi:10.1002/oby.22831.

15. NCD Risk Factor Collaboration (NCD-RisC). Worldwide trends in body-mass index, underweight, overweight, and obesity from 1975 to 2016: a pooled analysis of 2416 population-based measurement studies in 128.9 million children, adolescents, and adults. Lancet. 2017;390(10113):2627–2642. doi:10.1016/s0140-6736(17)32129-3.

16. GBD 2015 Obesity Collaborators. Health Effects of Overweight and Obesity in 195 Countries over 25 Years. N Engl J Med. 2017;377(1):13–27. doi:10.1056/nejmoa1614362.

17. Brasil. Ministério da Saúde. Vigitel Brasil 2018: vigilância de fatores de risco e proteção para doenças crônicas por inquérito telefônico. Brasília, DF: Ministério da Saúde; 2019. https://portalarquivos2.saude.gov.br/images/pdf/2019/julho/25/vigitel-brasil-2018.pdf. Accessed July 10, 2020.

18. GBD 2016 Brazil Collaborators. Burden of disease in Brazil, 1990-2016: a systematic subnational analysis for the Global Burden of Disease Study 2016. Lancet. 2018;392(10149):760–775. doi:10.1016/s0140-6736(18)31221-2.

19. Flint SW, Tahrani AA. COVID-19 and obesity-lack of clarity, guidance, and implications for care. Lancet Diabetes Endocrinol. 2020;8(6):474–475. doi:10.1016/s2213-8587(20)30156-x.

20. Brasil. Ministério da Saúde. Severe Acute Respiratory Syndrome dataset - including COVID-19 data. 2020. https://opendatasus.saude.gov.br/dataset/bd-srag-2020. Accessed July 11, 2020.

21. World Health Organization. Physical status: the use and interpretation of anthropometry. Geneva: WHO; 1995. https://apps.who.int/iris/handle/10665/37003. Accessed July 11, 2020.

22. Organización Panamericana de la Salud. División de Promoción y Protección de la Salud (HPP). Encuesta sobre Salud, Bienestar y Envejecimiento (SABE) en América Latina el Caribe: Informe Preliminar (Internet). In: XXXVI Reunión del Comité asesor de investigaciones em Salud; 9-11 jun 2001; Kingston: OPAS, 2002. http://envejecimiento.csic.es/documentos/documentos/paho-salud-01.pdf. Accessed July 11, 2020.

23. Brasil. Ministério da Saúde. Guidelines for the Collection and Analysis of Anthropometric Data in Health Services. Technical Standard for the Food and Nutrition Surveillance System. Brasília, DF: Ministério da Saúde; 2011.

24. Cheng FW, Gao X, Mitchell DC, et al. Body mass index and all-cause mortality among older adults. Obesity (Silver Spring). 2016;24(10):2232–39. doi:10.1002/oby.21612.

25. Honce R, Schultz-Cherry S. Impact of Obesity on Influenza A Virus Pathogenesis, Immune Response, and Evolution. Front Immunol. 2019;10:1071. doi:10.3389/fimmu.2019.01071.

26. Puig-Domingo M, Marazuela M, Giustina A. COVID-19 and endocrine diseases. A statement from the European Society of Endocrinology. Endocrine. 2020;68(1):2–5. doi:10.1007/s12020-020-02294-5.

27. Docherty AB, Harrison EM, Green CA, et al. Features of 20 133 UK patients in hospital with covid-19 using the ISARIC WHO Clinical Characterisation Protocol: prospective observational cohort study. BMJ. 2020;369:m1985. doi:10.1136/bmj.m1985.

28. Fernández-Niño JA, Guerra-Gómez JA, Idrovo AJ. Multimorbidity patterns among COVID-19 deaths: proposal for the construction of etiological models. Rev Panam Salud Publica. 2020;44:e166. doi:10.26633/RPSP.2020.166.

29. Williamson EJ, Walker AJ, Bhaskaran K, et al. Factors associated with COVID-19-related death using OpenSAFELY. Nature. 2020;584(7821):430–436. doi:10.1038/s41586-020-2521-4.

30. Klang E, Kassim G, Soffer S, et al. Morbid Obesity as an Independent Risk Factor for COVID-19 Mortality in Hospitalized Patients Younger than 50. Obesity (Silver Spring). 2020;28(9):1595–1599. doi:10.1002/oby.22913.

31. Cantarino L, Merchan-Hamann E. Influenza in Brazil: surveillance pathways. J Infect Dev Ctries. 2016;10(1):13–23. doi:10.3855/jidc.7135.

32. de Souza WM, Buss LF, Candido DDS, et al. Epidemiological and clinical characteristics of the COVID-19 epidemic in Brazil. Nat Hum Behav. 2020;4(8):856–865. doi:10.1038/s41562-020-0928-4.

33. Baqui P, Bica I, Marra V, Ercole A, van der Schaar M. Ethnic and regional variations in hospital mortality from COVID-19 in Brazil: a cross-sectional observational study. Lancet Glob Health. 2020;8(8):e1018–e1026. doi: 10.1016/S2214-109X(20)30285-0.

