## Supplementary Table 1 for "Combined association of obesity and other cardiometabolic diseases with severe COVID-19 outcomes: a nationwide cross-sectional study of 21,773 Brazilian adult and elderly inpatients"

**Suppl. Table 1**. Characteristics of the study population and study samples included and excluded of the analysis for each severe COVID-19 outcome.

| | Study population* | | Mechanical ventilation | | | | ICU admission | | | | Death | | | |
|---|---|---|---|---|---|---|---|---|---|---|---|---|---|---|
| | | | Included | | Excluded | | Included | | Excluded | | Included | | Excluded | |
| | n | % | n | % | n | % | n | % | n | % | n | % | n | % |
| Overall | 21,773 | 100.0 | 19,904 | 91.4 | 1,869 | 8.6 | 20,636 | 94.8 | 1,137 | 5.2 | 16,508 | 75.8 | 5,265 | 24.2 |
| **Sex** | | | | | | | | | | | | | | |
| Female | 9,742 | 44.7 | 8,905 | 44.7 | 837 | 44.8 | 9,227 | 44.7 | 515 | 45.3 | 7,379 | 44.7 | 2,363 | 44.9 |
| Male | 12,031 | 55.3 | 10,999 | 55.3 | 1,032 | 55.2 | 11,409 | 55.3 | 622 | 54.7 | 9,129 | 55.3 | 2,902 | 55.1 |
| **Age (years)** | | | | | | | | | | | | | | |
| 20-39 | 1,976 | 9.1 | 1,808 | 9.1 | 168 | 9.0 | 1,890 | 9.2 | 86 | 7.6 | 1,445 | 8.8 | 531 | 10.1 |
| 40-59 | 6,872 | 31.6 | 6,267 | 31.5 | 605 | 32.4 | 6,524 | 31.6 | 348 | 30.6 | 5,120 | 31.0 | 1,752 | 33.3 |
| 60-79 | 9,355 | 43.0 | 8,546 | 42.9 | 809 | 43.3 | 8,827 | 42.8 | 528 | 46.4 | 7,062 | 42.8 | 2,293 | 43.6 |
| >= 80 | 3,570 | 16.4 | 3,283 | 16.5 | 287 | 15.4 | 3,395 | 16.5 | 175 | 15.4 | 2,881 | 17.5 | 689 | 13.1 |
| **Obesity** | | | | | | | | | | | | | | |
| No | 20,463 | 94.0 | 18,661 | 93.8 | 1,802 | 96.4 | 19,366 | 93.9 | 1,097 | 96.5 | 15,533 | 94.1 | 4,930 | 93.6 |
| Yes | 1,310 | 6.0 | 1,243 | 6.2 | 67 | 3.6 | 1,270 | 6.2 | 40 | 3.5 | 975 | 5.9 | 335 | 6.4 |
| **Diabetes** | | | | | | | | | | | | | | |
| No | 13,058 | 60.0 | 11,925 | 59.9 | 1,133 | 60.6 | 12,401 | 60.1 | 657 | 57.8 | 9,925 | 60.1 | 3,133 | 59.5 |
| Yes | 8,715 | 40.0 | 7,979 | 40.1 | 736 | 39.4 | 8,235 | 39.9 | 480 | 42.2 | 6,583 | 39.9 | 2,132 | 40.5 |
| **Cardiovascular disease** | | | | | | | | | | | | | | |
| No | 10,391 | 47.7 | 9,400 | 47.2 | 991 | 53.0 | 9,858 | 47.8 | 533 | 46.9 | 7,866 | 47.7 | 2,525 | 48.0 |
| Yes | 11,382 | 52.3 | 10,504 | 52.8 | 878 | 47.0 | 10,778 | 52.2 | 604 | 53.1 | 8,642 | 52.4 | 2,740 | 52.0 |
| **Chronic pulmonary disease** | | | | | | | | | | | | | | |
| No | 20,387 | 93.6 | 18,630 | 93.6 | 1,757 | 94.0 | 19,306 | 93.6 | 1,081 | 95.1 | 15,422 | 93.4 | 4,965 | 94.3 |
| Yes | 1,386 | 6.4 | 1,274 | 6.4 | 112 | 6.0 | 1,330 | 6.5 | 56 | 4.9 | 1,086 | 6.6 | 300 | 5.7 |
| **Asthma** | | | | | | | | | | | | | | |
| No | 20,658 | 94.9 | 18,897 | 94.9 | 1,761 | 94.2 | 19,571 | 94.8 | 1,087 | 95.6 | 15,666 | 94.9 | 4,992 | 94.8 |
| Yes | 1,115 | 5.1 | 1,007 | 5.1 | 108 | 5.8 | 1,065 | 5.2 | 50 | 4.4 | 842 | 5.1 | 273 | 5.2 |
| **Chronic kidney disease** | | | | | | | | | | | | | | |
| No | 20,179 | 92.7 | 18,420 | 92.5 | 1,759 | 94.1 | 19,106 | 92.6 | 1,073 | 94.4 | 15,255 | 92.4 | 4,924 | 93.5 |
| Yes | 1,594 | 7.3 | 1,484 | 7.5 | 110 | 5.9 | 1,530 | 7.4 | 64 | 5.6 | 1,253 | 7.6 | 341 | 6.5 |
| **Chronic hematologic disease** | | | | | | | | | | | | | | |
| No | 21,438 | 98.5 | 19,597 | 98.5 | 1,841 | 98.5 | 20,313 | 98.4 | 1,125 | 98.9 | 16,245 | 98.4 | 5,193 | 98.6 |
| Yes | 335 | 1.5 | 307 | 1.5 | 28 | 1.5 | 323 | 1.6 | 12 | 1.1 | 263 | 1.6 | 72 | 1.4 |
| **Chronic neurological disease** | | | | | | | | | | | | | | |
| No | 20,459 | 94.0 | 18,700 | 94.0 | 1,759 | 94.1 | 19,377 | 93.9 | 1,082 | 95.2 | 15,460 | 93.7 | 4,999 | 95.0 |
| Yes | 1,314 | 6.0 | 1,204 | 6.1 | 110 | 5.9 | 1,259 | 6.1 | 55 | 4.8 | 1,048 | 6.4 | 266 | 5.1 |

| | | | | | | | | | | | | | | |
|---|---|---|---|---|---|---|---|---|---|---|---|---|---|---|
| **Chronic liver disease** | | | | | | | | | | | | | | |
| No | 21,418 | 98.4 | 19,581 | 98.4 | 1,837 | 98.3 | 20,300 | 98.4 | 1,118 | 98.3 | 16,218 | 98.2 | 5,200 | 98.8 |
| Yes | 355 | 1.6 | 323 | 1.6 | 32 | 1.7 | 336 | 1.6 | 19 | 1.7 | 290 | 1.8 | 65 | 1.2 |
| **Immunosuppression** | | | | | | | | | | | | | | |
| No | 20,579 | 94.5 | 18,792 | 94.4 | 1,787 | 95.6 | 19,485 | 94.4 | 1,094 | 96.2 | 15,571 | 94.3 | 5,008 | 95.1 |
| Yes | 1,194 | 5.5 | 1,112 | 5.6 | 82 | 4.4 | 1,151 | 5.6 | 43 | 3.8 | 937 | 5.7 | 257 | 4.9 |
